# Limited intestinal inflammation despite diarrhea, fecal viral RNA and SARS-CoV-2-specific IgA in patients with acute COVID-19

**DOI:** 10.1101/2020.09.03.20183947

**Authors:** Graham J. Britton, Alice Chen-Liaw, Francesca Cossarini, Alexandra E. Livanos, Matthew P. Spindler, Tamar Plitt, Joseph Eggers, Ilaria Mogno, Ana S. Gonzalez-Reiche, Sophia Siu, Michael Tankelevich, Lauren Tal Grinspan, Rebekah E. Dixon, Divya Jha, Adriana van de Guchte, Zenab Khan, Gustavo Martinez-Delgado, Fatima Amanat, Daisy A. Hoagland, Benjamin R. tenOever, Marla C. Dubinsky, Miriam Merad, Harm van Bakel, Florian Krammer, Gerold Bongers, Saurabh Mehandru, Jeremiah J. Faith

## Abstract

We sought to characterize the role of the gastrointestinal immune system in the pathogenesis of the inflammatory response associated with COVID-19. We measured cytokines, inflammatory markers, viral RNA, microbiome composition and antibody responses in stool from a cohort of 44 hospitalized COVID-19 patients. SARS-CoV-2 RNA was detected in stool of 41% of patients and more frequently in patients with diarrhea. Patients who survived had lower fecal viral RNA than those who died. Strains isolated from stool and nasopharynx of an individual were the same. Compared to uninfected controls, COVID-19 patients had higher fecal levels of IL-8 and lower levels of fecal IL-10. Stool IL-23 was higher in patients with more severe COVID-19 disease, and we found evidence of intestinal virus-specific IgA responses associated with more severe disease. We provide evidence for an ongoing humeral immune response to SARS-CoV-2 in the gastrointestinal tract, but little evidence of overt inflammation.

## Introduction

SARS-CoV-2 (Severe Acute Respiratory Syndrome CoronaVirus 2) a novel *Betacoronavirus* that causes COVID-19 (Coronavirus Disease 2019)) emerged in Wuhan, China December 2019 [1] and rapidly spread, leading to a worldwide pandemic. To date, over 30 million individuals worldwide have been infected and over 950,000 people have died (https://coronavirus.jhu.edu/map.html; accessed September 22 2020) [2].

COVID-19, primarily characterized by fever, cough and respiratory symptoms, is also now recognized to have a multitude of extrapulmonary manifestations [3] including gastrointestinal (GI) symptoms that are reported in up to 60% of hospitalized cases [3-5]. Intestinal inflammation in COVID-19 patients with diarrhea has been suggested by elevated levels of stool calprotectin, a protein released by neutrophils and used as a stool biomarker in inflammatory bowel disease (IBD) [6]. Furthermore, distinct alterations in the fecal microbiota are observed in COVID-19 patients [7], suggestive of immune dysregulation in the gut mucosa. Angiotensin-converting enzyme 2 (ACE2), the putative receptor for SARS-CoV-2, is highly expressed on the small intestinal epithelium [8, 9] and viral RNA has been detected in the stools of patients with COVID-19 for prolonged periods of time [10, 11]. Whether SARS-CoV-2 can actively replicate in the gut and be transmitted by this route is unknown, with conflicting reports in the literature [12-14]. SARS-CoV-2 can infect human-derived gut organoids (enteroids), supporting plausibility for infection of the gastrointestinal epithelium and a direct mucosal pathology that results in GI symptoms [15-17]. However, the exact pathophysiology of GI manifestations in COVID-19 is to be determined.

Systemic immune dysregulation is associated with severe COVID-19 infection, as high serum interleukin (IL)-6 IL-8, IL-10 and tumor necrosis factor (TNFα) correlate with increased disease severity and poor prognoses [18-23].

The role of the gastrointestinal tract in the pathogenesis of COVID-19 is not known but the intestinal immune system likely plays a crucial role in the inflammatory response following SARS-CoV-2 infection. Here, we sought to further characterize the GI manifestations in COVID-19 by examining intestinal virus, the fecal microbiome and intestinal immune responses in COVID-19 infected patients. We present results from a cohort of patients admitted to a large hospital in New York City during the peak of the COVID-19 pandemic.

## Results

### Patient demographics and clinical characteristics

We collected stool samples from 44 symptomatic COVID-19 patients hospitalized in New York City between April 15 and May 21 2020 (Table S1 and Date file S1). Samples were collected longitudinally during acute and convalescent phases of illness. The median time from symptom onset to sample collection of first stool sample was 16 days (range 2-66) and of second stool sample was 24.5 days (11-72) (Table S1 and Date file S2).

The majority of patients had at least one comorbidity (Table 1) and fifty percent of patients (n=22; 50%) presented to the hospital with moderate disease (see Methods for definitions of disease severity). During hospitalization 10 of the 22 (45.5%) patients who presented with moderate disease developed severe COVID-19 disease, while none of the patients who presented with mild symptoms had clinical worsening during hospitalization (Table 1 and Date file S1).

**Table 1:**
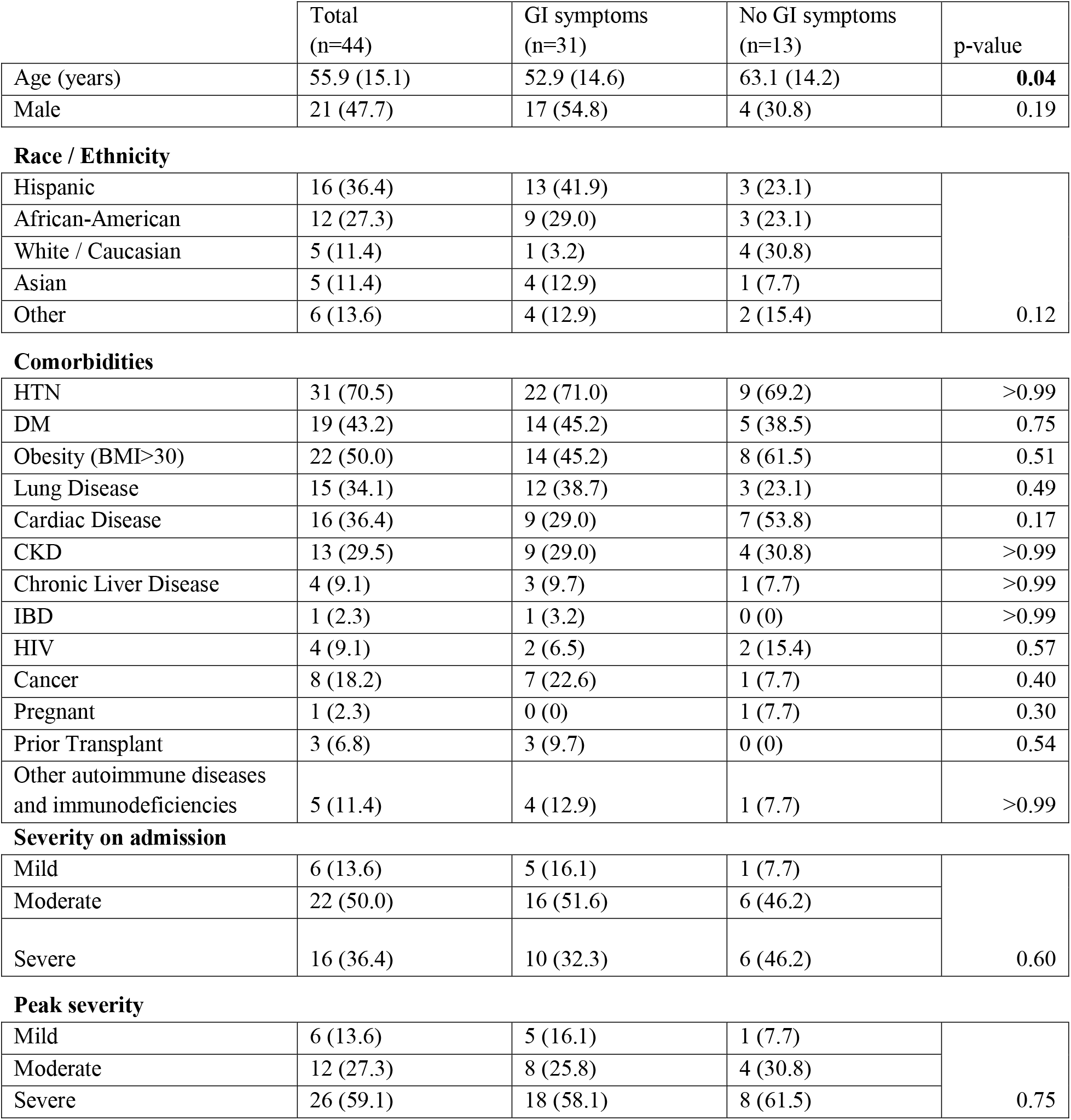
Patient Demographics and characteristics. The absolute number and percentage of the cohort (in parentheses) with each indicated characteristic. For Age, the median value (in years +/- the SD) is given. Statistical comparison of Ages is by Kruskal Wallis test and for other data by Fisher’s exact test.

|  | Total<br>(n=44) | GI symptoms<br>(n=31) | No GI symptoms<br>(n=13) | p-value |
| --- | --- | --- | --- | --- |
| Age (years) | 55.9 (15.1) | 52.9 (14.6) | 63.1 (14.2) | **0.04** |
| Male | 21 (47.7) | 17 (54.8) | 4 (30.8) | 0.19 |
| **Race / Ethnicity** |  |  |  |  |
| Hispanic | 16 (36.4) | 13 (41.9) | 3 (23.1) | 0.12 |
| African-American | 12 (27.3) | 9 (29.0) | 3 (23.1) |  |
| White / Caucasian | 5 (11.4) | 1 (3.2) | 4 (30.8) |  |
| Asian | 5 (11.4) | 4 (12.9) | 1 (7.7) |  |
| Other | 6 (13.6) | 4 (12.9) | 2 (15.4) |  |
| **Comorbidities** |  |  |  |  |
| HTN | 31 (70.5) | 22 (71.0) | 9 (69.2) | >0.99 |
| DM | 19 (43.2) | 14 (45.2) | 5 (38.5) | 0.75 |
| Obesity (BMI>30) | 22 (50.0) | 14 (45.2) | 8 (61.5) | 0.51 |
| Lung Disease | 15 (34.1) | 12 (38.7) | 3 (23.1) | 0.49 |
| Cardiac Disease | 16 (36.4) | 9 (29.0) | 7 (53.8) | 0.17 |
| CKD | 13 (29.5) | 9 (29.0) | 4 (30.8) | >0.99 |
| Chronic Liver Disease | 4 (9.1) | 3 (9.7) | 1 (7.7) | >0.99 |
| IBD | 1 (2.3) | 1 (3.2) | 0 (0) | >0.99 |
| HIV | 4 (9.1) | 2 (6.5) | 2 (15.4) | 0.57 |
| Cancer | 8 (18.2) | 7 (22.6) | 1 (7.7) | 0.40 |
| Pregnant | 1 (2.3) | 0 (0) | 1 (7.7) | 0.30 |
| Prior Transplant | 3 (6.8) | 3 (9.7) | 0 (0) | 0.54 |
| Other autoimmune diseases and immunodeficiencies | 5 (11.4) | 4 (12.9) | 1 (7.7) | >0.99 |
| **Severity on admission** |  |  |  |  |
| Mild | 6 (13.6) | 5 (16.1) | 1 (7.7) | 0.60 |
| Moderate | 22 (50.0) | 16 (51.6) | 6 (46.2) |  |
| Severe | 16 (36.4) | 10 (32.3) | 6 (46.2) |  |
| **Peak severity** |  |  |  |  |
| Mild | 6 (13.6) | 5 (16.1) | 1 (7.7) | 0.75 |
| Moderate | 12 (27.3) | 8 (25.8) | 4 (30.8) |  |
| Severe | 26 (59.1) | 18 (58.1) | 8 (61.5) |  |

Seven (15.9%) patients died and 13 patients (29.5%) were admitted to the intensive care unit (ICU) during their admission, with 14 patients (31.8%) meeting the composite outcome of ICU admission or death (Table 2). The majority of patients received antibiotics (28 (63.6%)) and therapeutic anticoagulation (31 (70.5%)) during their admission (Table S2 and Date file S1).

**Table 2:** Patient outcomes in patients with and without GI symptoms. The absolute number and percentage of the cohort (in parentheses) with each indicated outcome. Statistical comparisons are by Fisher’s exact test.

|  | Total<br>(n=44) | GI symptoms<br>(n=31) | No GI<br>symptoms<br>(n=13) | p-value |
| --- | --- | --- | --- | --- |
| Mortality | 7 (15.9) | 5 (16.1) | 2 (15.4) | >0.99 |
| ICU admission | 13 (29.5) | 6 (19.4) | 7 (53.8) | **0.03** |
| Composite outcome<br>(ICU admission or<br>death) | 14 (31.8) | 7 (22.6) | 7 (53.8) | 0.07 |

Given the aim of the study we prioritized enrollment of patients with GI symptoms. Thirty-one (70.5%) of patients in the cohort presented with GI symptoms (Figure 1A). The most reported GI symptom was diarrhea (59.1%), followed by nausea (34.1%) and then vomiting (15.9%) (Figure 1B). Patients with GI symptoms were significantly younger than patients without GI symptoms (mean (SD) age was 52.9 (14.6) vs 63.1 (14.2) respectively; p=0.04) (Table 1). Distribution of race and ethnicity and of comorbidities was similar in patients who presented with or without GI symptoms (Table 1). Ten (32.2%) of patients with and 6 (46.2%) without GI symptoms presented with severe disease. Overall distribution of disease severity on admission was similar in patients presenting with or without GI symptoms (p=0.60) as was peak severity during admission (p=0.75) (Table 1). Similar rates of mortality were observed in patients presenting with GI symptoms (n=5; 16.1%) compared to those without GI symptoms (n=2; 15.4%) (p > 0.99); however, significantly lower proportion of patients with GI symptoms (n=6; 19.4%) were admitted to the ICU compared with those without GI symptoms (n=7; 53.8%%, p=0.03; Fisher’s exact test) (Table 2). As expected, more severe disease at presentation (p=0.09) and more severe disease during admission (p=0.02), were associated with higher frequencies of ICU admission or death (Table S3).

**Fig. 1.**
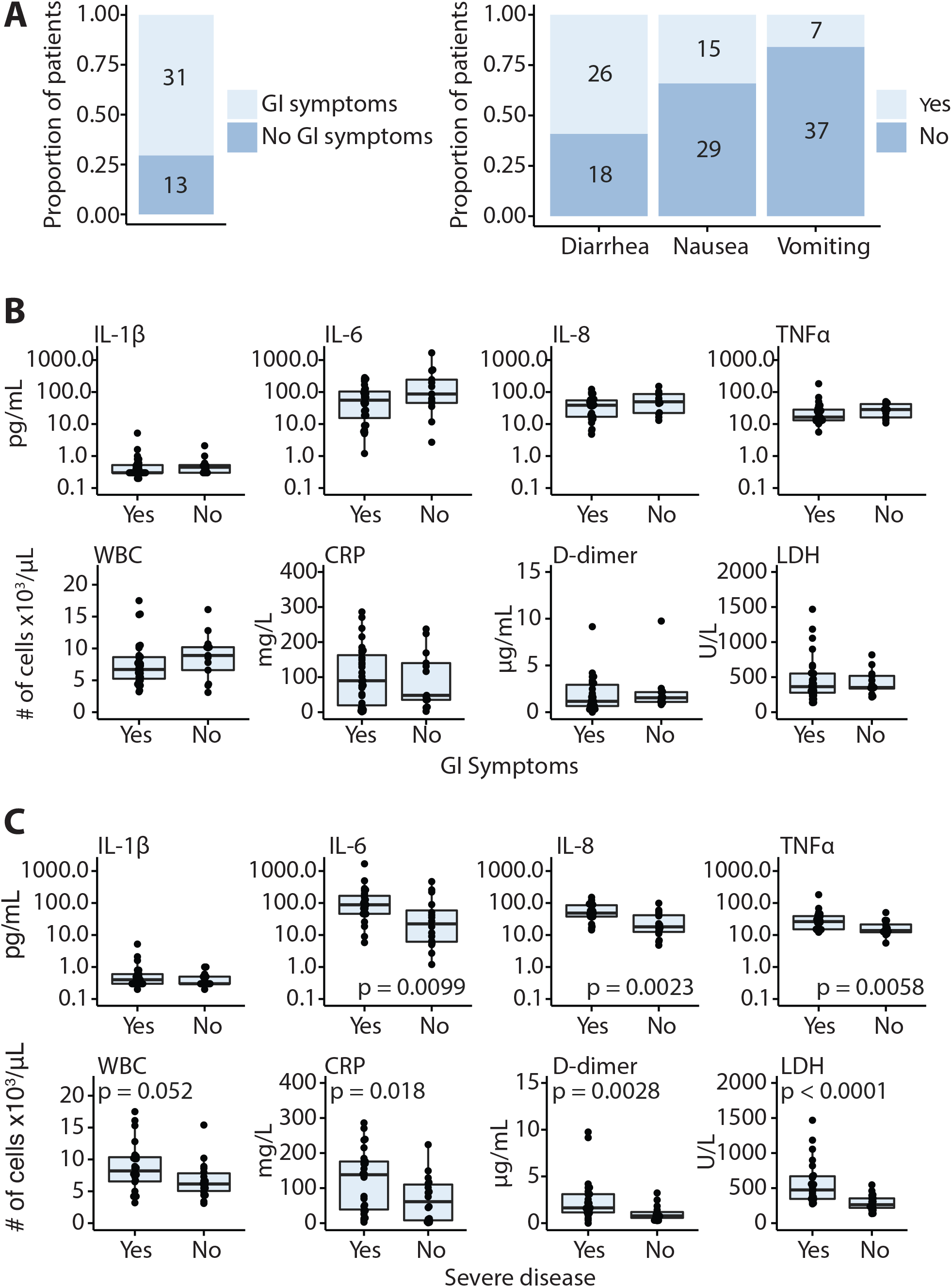
Gastrointestinal (GI) symptoms and serologic parameters in hospitalized COVID-19 patients. (**A**) Proportion of patients with GI manifestations in this cohort of hospitalized COVID-19 patients in whom stool samples obtained. **(B)** Serum concentrations of IL-1β, IL-6, IL-8, and TNFα and select laboratory values at the time of admission in patients with and without GI symptoms and **(C)** in patients with severe compared with non-severe COVID-19 disease. Each point represents an individual value for a patient, the box plot represents the median and the interquartile range and the p-values calculated using the Mann-Whitney test with significance defined as p<0.05.

When looking at inflammatory markers in serum, there were no significant differences observed between those with GI symptoms and those without, although those with GI symptoms tended to have lower serum IL-6 and IL-8 (Fig. 1B). No notable differences in serum inflammatory markers or laboratory values were noted in patients with GI symptoms or with severe disease (Fig. 1B, S1A and B).

Peak disease severity was associated with significantly higher serum IL-6, IL-8, and TNF-α consistent with prior reports [19, 20, 23] (Fig. 1C). Additionally, severe disease was associated with higher serum C-reactive protein (CRP), Lactate dehydrogenase (LDH), D-dimer, ferritin and procalcitonin (Fig. 1C and Fig. S1B). There were no significant differences in liver enzymes in those with severe disease compared to those with mild or moderate disease (Fig. S1B).

### SARS-CoV-2 RNA is frequently detected in stool and higher viral loads are associated with diarrhea and death

To detect the presence of SARS-CoV-2 RNA in stool, we performed real-time quantitative PCR (qPCR) targeting three regions of the SARS-CoV-2 genome (see Methods for details). A positive result was defined as two out of three SARS-CoV-2 primers with cycle thresholds (Ct) <40. Eighteen of the 44 patients (40.9%) had/showed a positive fecal PCR over this threshold during admission (21 of 62 samples tested (33.9%)).

To investigate the length of time SARS-CoV-2 RNA may be found in the stool, we stratified our qPCR results by the time from symptom onset. SARS-CoV-2 RNA was unlikely to be detected in stool samples more than 28 days after symptom onset (p = 0.052; Fig. 2A-B and S2A). Only 2/17 (11.8 %) tests were positive for viral RNA in the stool after 28 days from symptom onset as opposed to 19/45 (42.2%) of tests before this point (p=0.03, Fisher exact test; Fig. 2B and S2A). However, within the initial 28-day period there was no difference as to when stool samples tested positive for viral RNA (p = 0.24; Fig. 2A and S2A). Interestingly, we never detected viral genome in stool collected after a patient had a recorded negative nasopharyngeal PCR result (n=7 stool samples from 5 different patients collected after a negative nasal PCR; median 11 days between negative nasal PCR and stool sample collection in this subset).

**Fig. 2.**
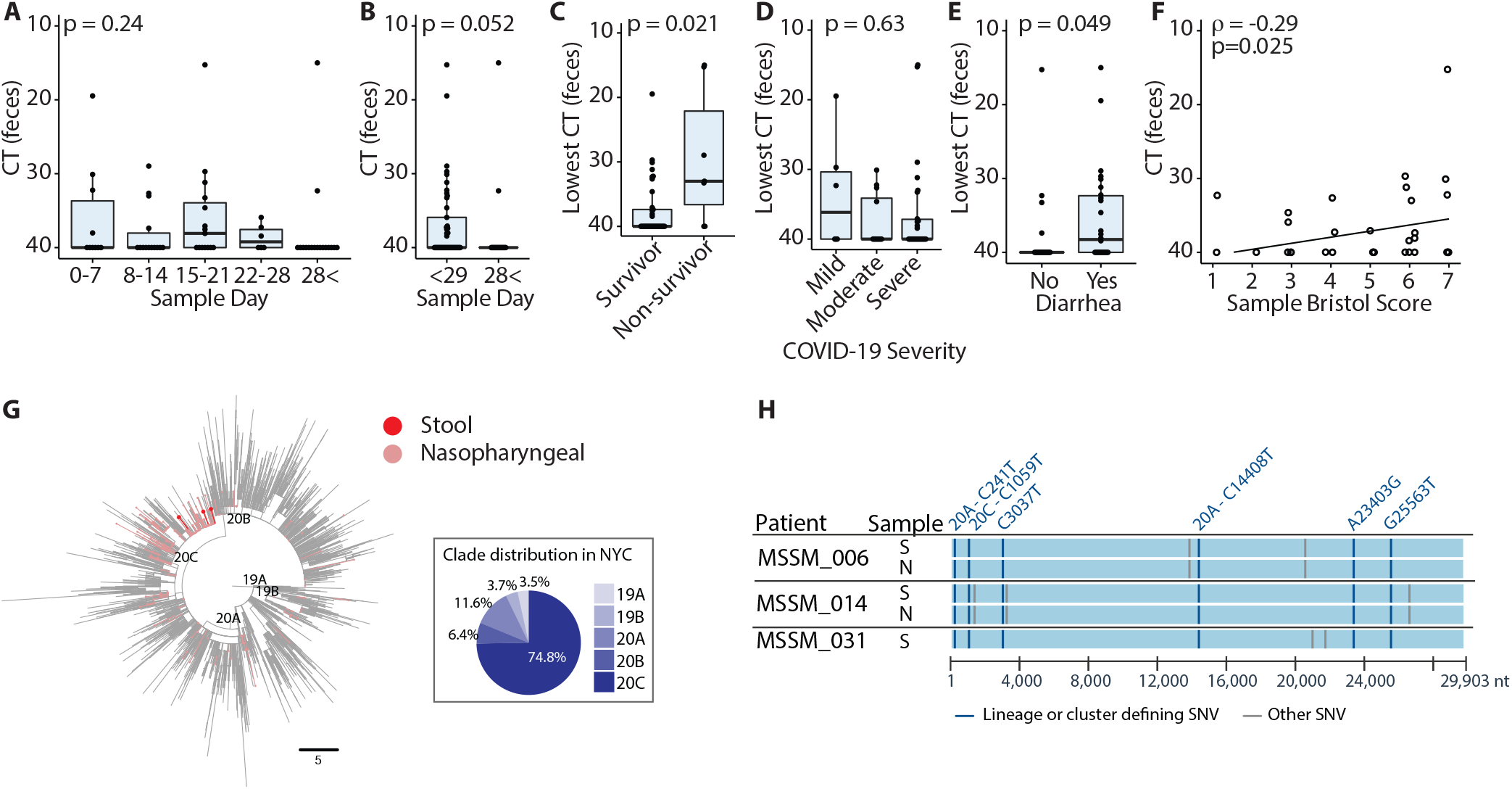
Detection and sequencing of SARS-CoV-2 virus genome in stool. **(A)**. The median fecal SARS-CoV-2 Ct of each sample, plotted according to the time of collection relative to either the onset of symptoms or the first positive nasal PCR. P value - Kruskal-Wallis. (**B)**. The median fecal SARS-CoV-2 Ct of each sample, plotted according to the relative time of collection: day 28 as a cut off. Relative to either the onset of symptoms or the first positive nasal PCR. P value - Mann-Whitney. (**C)**. The lowest fecal SARS-CoV-2 Ct measured for each donor shows higher peak viral loads in donors who did not survive. P value - Mann-Whitney. (**D)**. The lowest fecal SARS-CoV-2 Ct value measured from each donor during admission, plotted according to the peak severity of COVID19 symptoms- P value - Kruskal-Wallis. **(E)**. Fecal SARS-CoV-2 Ct values in samples from donors with and without diarrhea. P value - Mann-Whitney. (**F**). The relationship between the fecal SARS-CoV-2 Ct values and the Bristol Score of each sample. Spearman rank test. (**G**) Phylogenetic relationships of the SARS-CoV-2 isolates from stool (Stool, dark red) and other nasopharyngeal virus isolates from the Mount Sinai Health System (MSHS) in NYC (Nasopharyngeal, salmon), in a background of global strains (grey). Main phylogenetic clades are indicated. The scale bar represents divergence from the root (reference genome NC_045512) as number of single nucleotide variants (SNVs). The panel in the bottom right indicates the clade distribution of the MSHS nasopharyngeal samples from 403 SARS-CoV-2 genomes from February 29 to July 15. (**H**) Sequence alignment depicting clade and cluster defining (teal), and other (grey) SNVs present in the stool SARS-CoV-2 isolates from stool (S) or nasopharynx (N) of the indicated donors. The clade and amino acid substitutions are indicated when applicable.

To determine whether fecal viral genome loads were associated with COVID-19 symptoms or outcomes, we stratified our results and found that patients who did not survive had significantly higher fecal SARS-CoV-2 genome loads than survivors (p = 0.021; Fig. 2C). However, there was no association between the fecal Ct values and disease severity (p = 0.63; Fig. 2D), and it was notable that 3/6 (50%) of patients with mild COVID-19 disease had detectable virus in stool (Fig. 2D). We observed higher viral genome loads in samples from patients reporting diarrhea (p 0.049; Fig. 2E and S2B) and a significant correlation between viral genome load and Bristol stool scores of the samples (Spearman ρ = −0.29, p = 0.025; Fig. 2F).

We sequenced the genome of SARS-CoV-2 genomes from stool samples of three patients using short read sequencing [24]. Each of these isolates belonged to clade 20C, the clade also most commonly isolated from nasopharyngeal samples in New York City during this period of time. The mutation profiles of these samples suggest that there were no unusual substitutions particularly associated with GI infection (Fig. 2G and H). From two of these patients we also obtained SARS-CoV-2 genomes isolated from the nasopharynx. These strains were identical to those isolated from stool of the same patient (Fig. 2H).

### Fecal inflammatory cytokines are elevated in COVID-19 cases but are unrelated to gastrointestinal symptoms

The detection of SARS-CoV-2 genome in the stool of multiple donors and the inverse association with fecal consistency led us to investigate if viral infection was causing intestinal inflammation associated with diarrhea among this cohort. Cytokines can be measured in fecal samples, and this method has been used as a sensitive and non-invasive way to monitor intestinal immune responses in inflammatory disease and during enteric infection [25-31]. In particular, fecal levels of IL-6, IL-8 and IL-1β are elevated in the context of acute bacterial or viral gastroenteritis and ulcerative colitis [28, 29]. We measured concentrations of eight cytokines and calprotectin in the stool samples from the COVID-19 cohort and in a group of stool samples collected from healthy donors recruited before SARS-CoV-2 was endemic in New York City (see Methods for details).

We first examined differences in fecal cytokines between COVID-19 patients and uninfected controls. Levels of fecal IL-8 were significantly elevated while levels of IL-10 were significantly lower in COVID-19 patients compared to uninfected controls (Fig. 3A), while the remaining tested cytokines were not significantly different. Although we observed elevated levels of IL-1β and TNFα in some COVID-19 patients, this was not consistent across the cohort (Fig. 3A). Next, we analyzed fecal cytokines in COVID-19 patients stratified by disease severity. IL-23 was the only cytokine that was significantly different between groups with higher a concentration in patients with severe COVID-19 disease (Fig. 3B). The remainder of fecal cytokines analyzed (IL-1β, IL-1ra, IL-6, IL-8, IL-17, TNFα) were not significantly different in COVID-19 patients compared to controls nor associated with severity of disease (Fig. 3A and B). We found no difference in the concentration of any fecal cytokine between those patients reporting diarrhea compared to those without diarrhea (Fig. S3A). Fecal concentrations of IL-1β, IL-6, IL-8and TNFα were not associated with concentrations of these cytokines measured in serum at the time of admission, although it is important to note that these samples were collected on different study days (median 16 days, range 2-66 days between samples; Fig. S3B). Fecal calprotectin, a marker of intestinal inflammation, was not associated with COVID-19 disease severity (p=0.12; Fig. 3C) nor stool viral genome load (p=0.22; Fig. 3D).

**Fig. 3.**
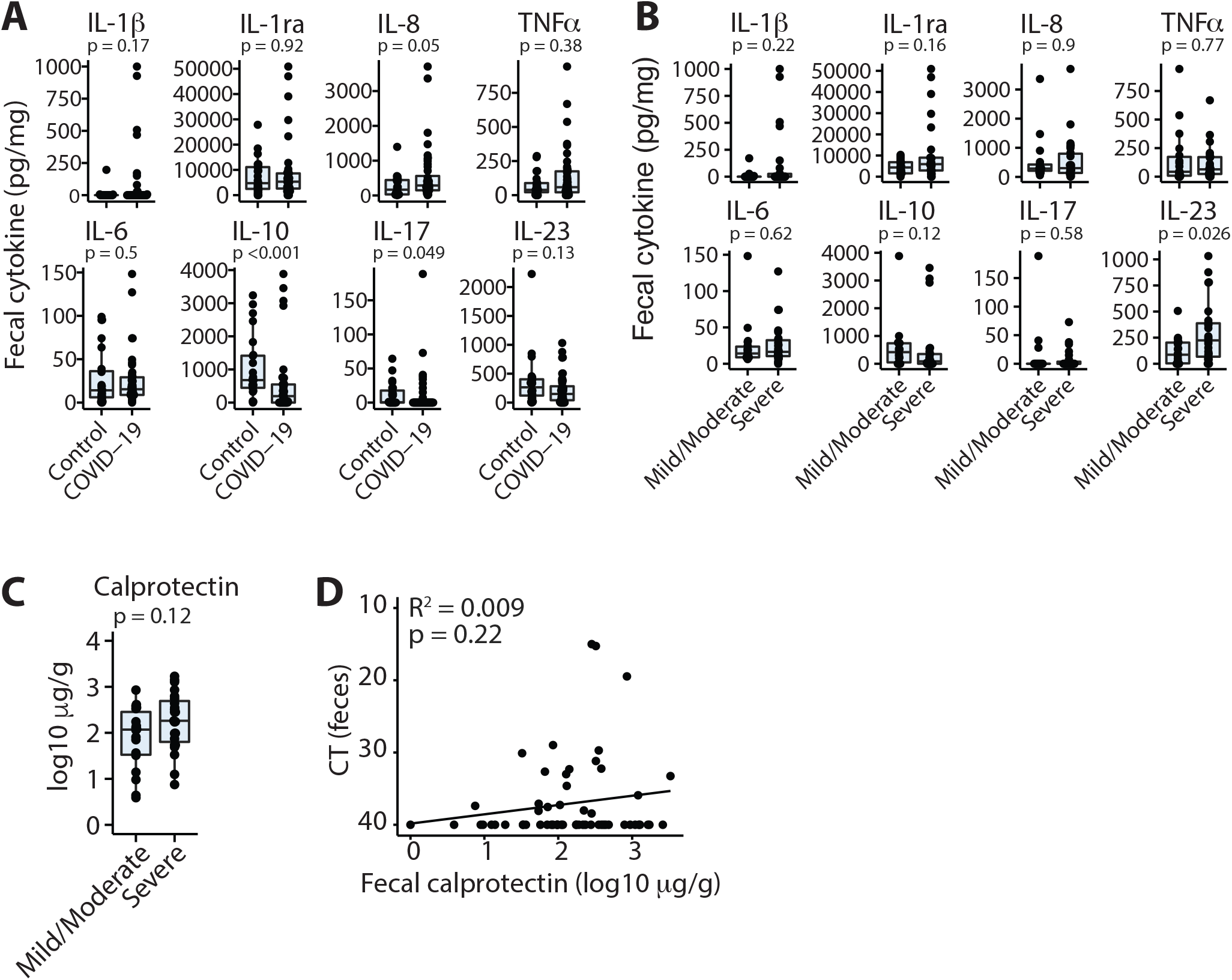
Fecal cytokine levels in COVID-19 patients. **(A)**. Concentrations of cytokines in fecal samples from control donors and COVID-19 patients. Where multiple samples from the same donor were collected, the median of these samples is plotted. (**B)**. Concentrations of fecal cytokines in COVID-19 patients with severe disease compared to those with non-severe disease. **(C)** Concentrations of fecal calprotectin in COVID-19 patients with severe disease compared to those with non-severe disease. **(D)** Concentrations of fecal calprotectin are unrelated to fecal viral load. Boxplots show median +/- IQR and P values are calculated by Mann-Whitney test.

### The gut microbiome of COVID-19 patients is altered by antibiotic exposure but unrelated to disease severity or GI symptoms

A common feature of intestinal inflammation is disruption to the structure and diversity of the gut microbiota (dysbiosis) and there have been reports of gut microbiota changes in COVID-19 patients [7]. We evaluated the fecal microbiome of the COVID-19 patient cohort by both metagenomic shotgun and 16S rRNA gene amplicon sequencing. Analysis of both datasets revealed no features of composition, structure or diversity that associated with disease severity (Fig. 4A). Furthermore, we found no consistent changes in gut microbiome composition or richness in samples from patients reporting diarrhea, although the Shannon entropy, a metric of community richness and evenness, was slightly lower in samples with higher Bristol scores (p=0.034; Fig. 4B). Patients currently or recently treated with antibiotics, particularly vancomycin and/or ceftriaxone showed significant compositional disruption and reduced diversity as measured by both 16S (p=0.03) and metagenomic (p=0.0079) sequencing (Fig. 4C). These data further support a relative lack of severe inflammation, which would be expected to skew the composition of the microbiota, and suggests that antibiotic therapy, common among hospitalized COVID-19 patients, has a greater impact on the gut microbiome than SARS-CoV-2 infection itself, regardless of GI symptoms or clinical course.

**Fig. 4.**
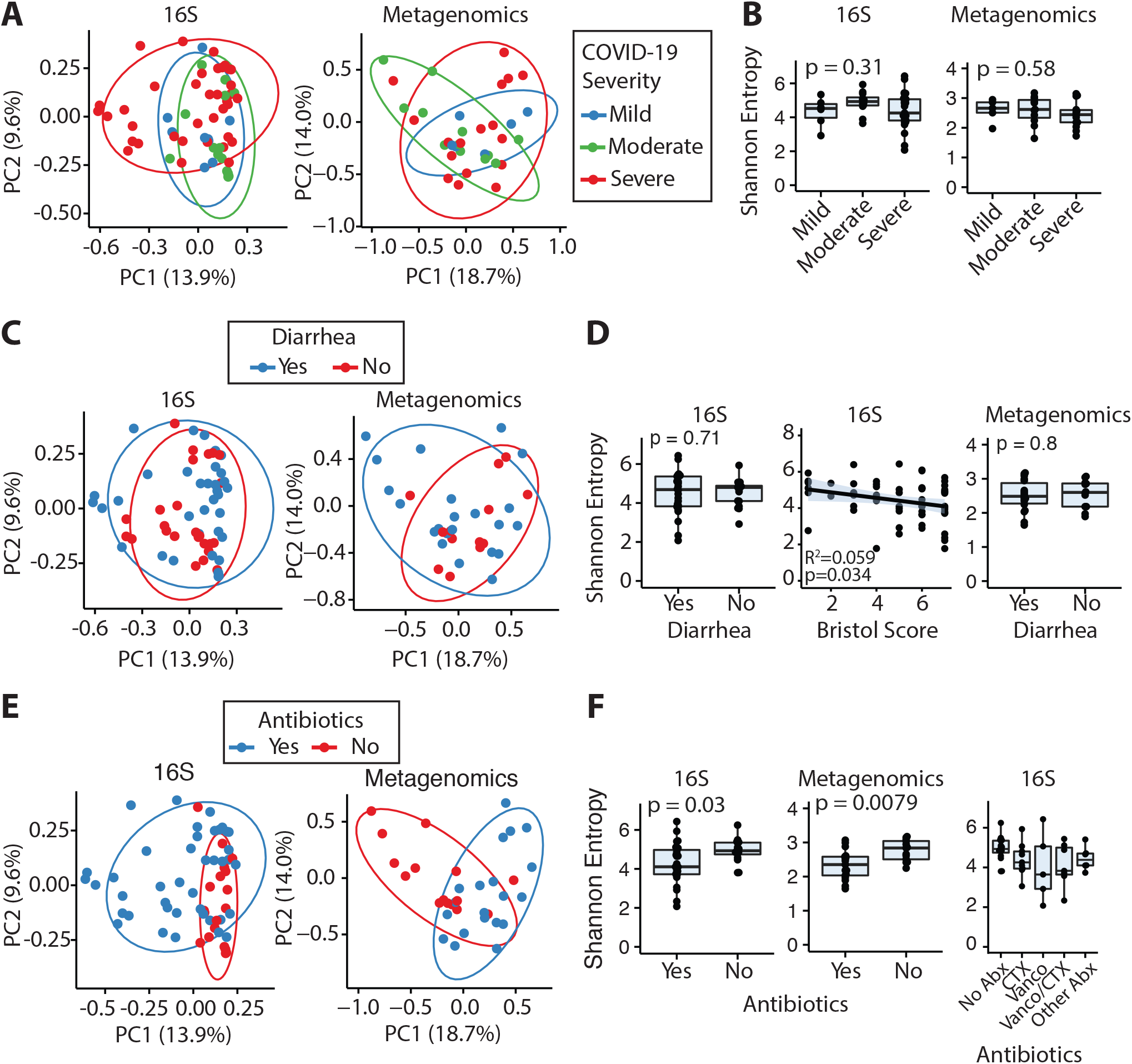
Gut microbiome and fecal SARS-CoV-2 genome sequencing of COVID-19 patients. **(A)** PCoA (Bray-Curtis dissimilarity) of 16S rRNA gene amplicon and shotgun metagenomic sequencing of fecal samples from donors stratified by COVID-19 severity. (**B**) Alpha diversity (Shannon entropy) is not significantly altered in mild, moderate or severe COVID-19. **(C)** PCoA (Bray-Curtis dissimilarity) of 16S rRNA gene amplicon and shotgun metagenomic sequencing of fecal samples from donors with and without diarrhea. (**D**) Gut microbiome alpha diversity (Shannon entropy) in patients with and without diarrhea and as relates to the Bristol score of each sample. **(E)** PCoA (Bray-Curtis dissimilarity) of 16S rRNA gene amplicon and shotgun metagenomic sequencing of fecal samples from patients receiving or not receiving antibiotic therapy. (**F**). Gut microbiome alpha diversity (Shannon entropy) is reduced in hospitalized COVID-19 patients recently or currently receiving antibiotic therapy (Abx). Boxplots show median +/- IQR.

### SARS-CoV-2 specific IgA is found in the stool of patients with severe COVID-19

A major component of the immune response against SARS-CoV-2 is believed to be the development of neutralizing antibodies against the viral Spike (S) protein, specifically the receptor binding domain (RBD) [32]. IgA is the major mucosal isotype and is known to provide long-term protection from enteric viruses, including rotavirus [33]. SARS-CoV-2-specific IgA has been detected in serum [32] and is found secreted in bronchoalveolar lavage fluid, nasal washes and in breast milk of convalescent donors [34-36]. We sought to determine if individuals with COVID-19 mounted an analogous gut mucosal IgA response against SARS-CoV-2, and if this was related to fecal viral load or other aspects of their clinical course.

We modified a direct enzyme linked immunosorbent assays (ELISA) assay developed to detect the presence of anti-RBD antibodies in the serum [32, 37] to measure fecal RBD-specific IgA (see Methods for details). Stool samples were treated with 0.5% Triton-X 100 for 1 hour to inactivate potentially infectious virus. Importantly, we found Triton-X 100 did not interfere with the detection of RBD-specific IgA (Fig. S4A).

The level of fecal RBD-reactive IgA was correlated with serum RBD-specific levels (R^2^=0.14, p=0.026, f-test; Fig. 5A). However, titers in feces were lower than in serum (Fig. 5A and S4B) and in contrast to serum we observed significant background signal at higher fecal concentrations, even among samples from healthy uninfected controls, which we attribute to non-specific binding of proteins from the high-complexity fecal samples (Fig. 5B). Across the cohort, there was no significant difference in the level of RBD-specific IgA detected in stool in healthy controls and COVID-19 patients (p=0.48, Mann-Whitney test; Fig. 5C), nor in the total amount of fecal IgA (p=0.76, Mann-Whitney test; Fig. S4C). Five out of forty-four COVID-19 patients (11%) demonstrated substantial fecal IgA reactivity to the SARS-CoV-2 RBD above the background level (AUC >3.5; Fig. 5B and 5C). The 5 patients with detectable levels of fecal RBD-specific IgA were all among those with severe COVID-19 (Fig. 5D) and notably, all these subjects were obese (median BMI of 40.7 vs 28.8 for those without detectable levels, p=0.007; Mann-Whitney test).

**Fig. 5.**
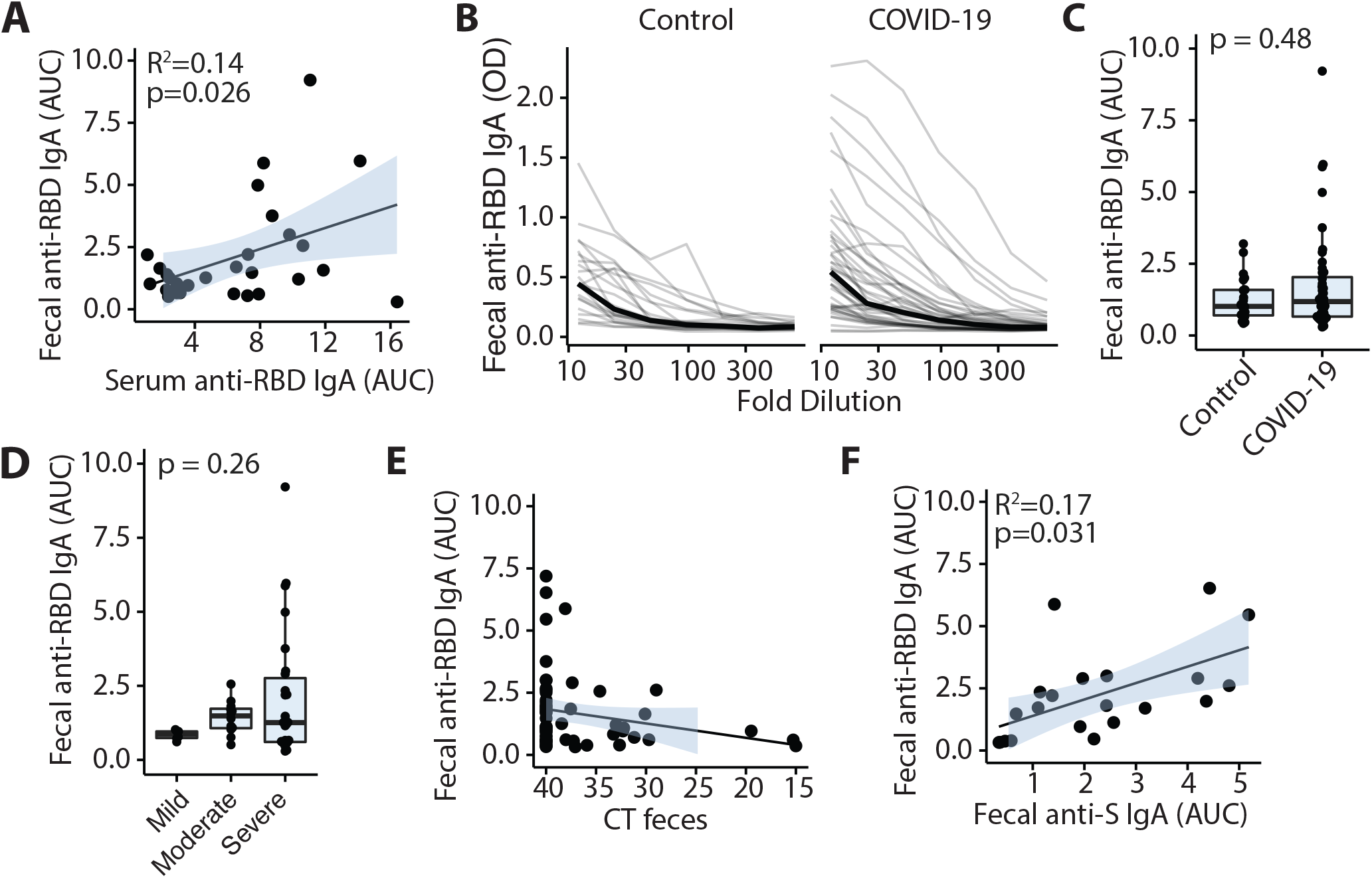
SARS-CoV-2-specific IgA in stool of COVID-19 patients. **(A)**. Fecal and serum anti-SARS-CoV-2 RBD IgA are correlated. **(B**,**C)**. Relative titers of anti-SARS-CoV-2 S1 RBD IgA in feces of control and COVID infected individuals. (**D)**. Relative titers of anti-SARS-CoV-2 S1 RBD IgA in feces of COVID infected individuals with varying disease severity. **(E)**. Anti-SARS-CoV-2 S1 RBD IgA and viral load are not correlated. Each point shows data from a single sample. **(F)**. Anti-SARS-CoV-2 S1 RBD IgA and anti-S1 IgA in feces are correlated. Each point shows data from a single sample.

Of those patients with detectable fecal RBD-specific IgA (AUC > 3.5), 4/5 (80%) reported diarrhea, but across the whole cohort fecal RBD-specific IgA was not associated with diarrhea, Bristol stool score or the level of viral RNA in feces (p=0.89, Wilcox test; Fig. 5E Fig. S4D and S4E). We sampled COVID-19 patients longitudinally and found an individual’s fecal IgA reactivity to RBD was relatively stable over the sampling period – up to 30 days for some donors (Fig. S4F), similar to what is observed in systemic circulation[38]. Additionally, a subset of samples was tested for fecal spike protein IgA reactivity which was strongly correlated with fecal RBD reactivity (Fig. 5F). As with fecal RBD specific IgA, we did not observe significantly different spike-specific fecal IgA between COVID-19 and control donors (Fig. S4H).

## Discussion

We describe analysis of fecal samples from a cohort of 44 patients with COVID-19 where we sought to characterize the nature of the intestinal immune responses to SARS-CoV-2 and investigate immunological associations with GI symptoms. The cohort comprised exclusively hospitalized patients and patients with GI symptoms were prioritized for recruitment. In approximately 40% of individuals, we detected SARS-CoV-2 genome in feces by qPCR, with higher viral loads among donors with diarrhea, consistent with a recent report of hospitalized patients in Hong Kong [5]. Although detection of viral RNA does not prove infectivity, these results point towards productive SARS-CoV-2 infection of the intestines. This is supported by expression of ACE2, the receptor for SARS-CoV-2 in the intestines [14, 39, 40] and by data showing that human gut enteroids support SARS-CoV-2 infection [17]. Diarrhea in setting of viral infection can be inflammatory or non-inflammatory in nature, given lack of correlation with fecal calprotectin, this data suggest that diarrhea is a result of non-inflammatory mechanism and this finding might warrant further exploration in future studies.

Virus genome was detected in stool of 50% of those with mild COVID-19 symptoms but fecal viral RNA was also higher among patients who did not survive. This likely represents two distinct things; a milder disease course is associated with GI symptoms [41], and complex immune dysregulation in severe cases may lead to uncontrolled viral replication in multiple organs [42]. Strains isolated from stool were the same as isolates from the nasopharynx of the same patient and all stool isolates belonged to the clade most prevalent in New York City. This suggests that GI infection is not associated with any specific clade or strain.

The higher fecal IL-8 and lower IL-10 concentration in COVID-19 patients compared to controls and the higher IL-23 in patients with severe disease point to some degree of immunological involvement of the GI tract in SARS-CoV-2 infection. However, despite frequent GI symptoms and the presence of viral RNA in stool, this response was generally mild, given that no other fecal cytokines or calprotectin were found to be elevated in patients with COVID-19.

Change in the structure and composition of the intestinal microbiome is also a marker of intestinal inflammation. The changes in the intestinal microbiome that we observed were mostly driven by the use of antibiotics rather than the severity of COVID-19 infection or the presence of diarrhea. These findings support the conclusion that intestinal SARS-CoV-2 infection elicits a mild inflammatory response in the gut.

We detected anti-SARS-CoV-2 antibody responses in the stool, presenting the first evidence of RBD-specific IgA in fecal samples from COVID-19 patients. We found that the fecal RBD-specific IgA response correlated with serum RBD-specific IgA response. It remains to be determined whether fecal SARS-CoV-2 specific IgA represents a localized protective response within the intestines or reflects a systemic response to the viral challenge. The ability to detect specific IgA in feces is likely more challenging in stool than in serum due reduced signal-to-noise, and low levels of specific fecal antibody if present in some patients would not be detected with our assay. There is evidence that secretory IgA is highly neutralizing, further supporting the potential importance of the mucosal immune response in COVID-19 disease [43].

In summary, our data suggests that the gut can be an immunologically active organ during SARS-CoV-2 infection, as evidenced by virus-specific IgA, but there is little evidence for overt intestinal inflammation, even in patients with diarrhea or other GI symptoms.

## Materials and Methods

### Patient recruitment

We daily screened all consecutive patients admitted to the Mount Sinai Hospital for COVID-19 infection from April 15 2020 to May 21 2020. The study was reviewed and approved by the institutional review board at our medical center (Protocol #: HS# 16-00512/ GCO# 16-0583). We enrolled patients who were able to give informed consent and were able to provide a stool sample. We excluded patients who were not able to provide consent at the time of SARS-CoV-2 testing (sedation, endotracheal intubation prior or at the time of Emergency Room arrival, dementia or any other condition that prevented the subject to give fully informed consent). As our focus was to evaluate the role of gastrointestinal (GI) symptoms in the course of disease, we actively enrolled all patients with documented diarrhea, nausea and / or vomiting on presentation. We also enrolled patients without GI symptoms presenting at the same time to have a comparison group. We enrolled a total of 66 patients. Three patients were asymptomatic from SARS-CoV-2 infection and were excluded. Of the remaining 63, we collected stool samples from 44 and only these patients are considered in these analyses.

### Clinical data collection; questionnaire and chart review

A questionnaire regarding symptoms onset and type of symptoms experienced was administered to all subjects. Detailed questions regarding gastrointestinal symptoms were included in the questionnaire (Supplemental Item 1). Chart review was used to collect subjects’ demographic characteristics (gender, age, race and ethnicity), comorbidities and medical history, COVID-19 clinical presentation, course of disease, laboratory data and COVID-19 treatment. Subject medical history was reviewed, and relevant comorbidities (hypertension, diabetes mellitus, obesity, chronic kidney disease, underlying chronic lung or cardiac diseases, inflammatory bowel disease, chronic HIV-infection, autoimmune diseases, current or prior history of cancer, current or prior history of organ transplant, concomitant pregnancy) were recorded.

COVID-19 disease severity was defined based on our institutional algorithm. Mild disease was defined as having a peripheral oxygen saturation (O2 Sat) ≥ 94% and no evidence of pneumonia on chest radiographic imaging (CXR); moderate disease was defined as either evidence of pneumonia on CXR or O2 Sat < 94%; severe disease was defined as evidence of pneumonia on CXR and the need for use of supplemental oxygen in the form of non-rebreather mask, high flow nasal cannula or mechanical ventilation. Severe disease was further subclassified as with or without evidence of end organ damage defined as: need for vasopressors support, creatinine clearance < 30 mL/min, alanine aminotransferase (ALT) > 5 times the upper limit of normal. Disease severity was recorded at the time of admission. Subjects were classified based on their symptoms at presentation in patients with and without GI symptoms. Qualifying GI symptoms were diarrhea, nausea or vomiting (at least one episode of self-reported symptom per day prior to presentation). Date of overall symptoms and of GI symptoms were also recorded. Clinical outcomes were collected as maximum level of oxygenation required during hospitalization (room air, nasal cannula, non-rebreather mask, high flow nasal cannula or mechanical ventilation), need for Intensive Care Unit (ICU) admission and death or discharge from the hospital.

Laboratory data were also collected via chart review, we included results for: SARS-CoV-2 nasal PCR, SARS-CoV-2 antibodies, peripheral WBC and lymphocyte percentage, Aspartate Aminotransferase (AST), Alanine Aminotransferase (ALT), Alkaline Phosphatase (ALKP), C-Reactive Protein, Procalcitonin, Lactic Acid Dehydrogenase (LDH), Ferritin, D-Dimer and cytokines that were implemented as part of clinical care IL-6, IL-8, TNFα and IL-1β. Serum cytokines were measured using the Ella platform as previously described[23]. Laboratory values were collected at baseline (at the time of admission) and at the time of discharge or death. Peak values for each lab during the admission was also collected.

COVID-19 treatment data was also collected, including use of hydroxychloroquine, azithromycin, systemic anticoagulation, steroids, convalescent plasma, and remdesivir. Information regarding subject’s participation on double blinded clinical trials was recorded. Use of antibiotics and class of antibiotics was also captured. All deidentified clinical data is provided in Data File S1.

Fecal samples from 22 healthy donors were previously collected from donors recruited in New York City between 2015 and 2016 as part of an unrelated study reviewed by the Mount Sinai Institutional Review Board (HS# 11-01669) and were stored at –80C until analysis. The mean (SD) age in the control group was 52.1 (11.3) compared to 55.9 (15.1) of the COVID-19 cohort (p=0.3, t-test) and 9 healthy donors were male (40.9%) compared with 47.7% of the COVID-19 cohort.

### Stool sample collection and processing

Stool donations were collected into sterile containers (Covidien Precision™ Stool Collector) and kept at 4°C for no more than 24 hours before processing. Working under aseptic conditions in a BSL2+ laboratory, the sample was subdivided into aliquots for subsequent analyses. For the measurement of antibodies and cytokines, stool aliquots of 150-600 mg were frozen at −80°C. Samples were thawed on ice and homogenized to a concentration of 500 mg/mL in 1x PBS supplemented with protease inhibitor cocktail (cOmplete EDTA-free, Roche (1 tablet in 10mL) and phenylmethylsulfonyl fluoride (0.3 mM); to prevent protein degradation) and Triton X-100 (0.5% final concentration; to inactivate SARS-CoV-2 virus [44]). Samples were homogenized by vigorous shaking in a Mini-Beadbeater (Biospec) without beads for 60 seconds, kept on ice for one hour to ensure virus inactivation, centrifuged at 12,000 x g for 20 minutes at 4°C and the supernatants collected. For measurement of fecal calprotectin, stool aliquots of 150 - 600 mg were placed into 1 mL PBS containing protease inhibitor cocktail (cOmplete EDTA-free, Roche; 1 tablet in 10mL), placed at 60°C for 1 hour to inactivate SARS-CoV-2 before freezing at −80°C. For SARS-CoV-2 qPCR, stool aliquots (89.2 mg ± 77.8 (SD)) were immediately placed into 0.5 mL of Trizol and homogenized in a Mini-Beadbeater for 2 minutes. The mass of stool used for extraction did not affect positivity rate (p=0.3, Mann-Whitney test). The samples were stored at −80°C until RNA extraction. For culture independent gut microbiome sequencing, stool aliquots were immediately added to 1.3 mL of extraction buffer (0.55 mL 25:24:1 Phenol:Chloroform:Isoamylalcohol, 0.28 mL 2x STE Buffer (20mM Tris, 2mM EDTA, 2mM NaCl, pH 8), 0.27 mL QIAquick PM buffer (Qiagen) and 0.2 mL 20% SDS) and bead beat for 2 minutes with ∼400ul of 0.1mm silica beads. After centrifugation (8000rpm for 1 minute), the aqueous phase was removed and stored at −80°C for additional purification as described below.

### Blood sample collection and processing

Research study blood samples were collected with the next clinically indicated blood draw after enrollment and were processed within four hours of collection. Serum was collected in a Tiger-top tube (serum separator tube) and stored directly at −80 degrees after being spun.

### SARS-CoV-2 PCR from fecal samples

#### RNA extraction

Total RNA was extracted from stool samples by Trizol™ (Thermo Fisher, Cat. No. 15596026) and the PureLink™ RNA Mini Kit (Thermo Fisher, Cat. No. 12183020). After the frozen stool samples in Trizol were thawed, an additional 0.5 mL of Trizol was added to each tube for a total of 1 mL of Trizol per stool sample. The samples were mixed gently by inverting 5 times and centrifuged at 12000g for 3 minutes at 4°C to remove any remaining stool debris. Following centrifugation, ∼1 mL of the supernatant (containing RNA) was transferred to a new 2 mL tube and 0.2 mL of chloroform was added to each tube. The samples were mixed by inverting vigorously until the mixture was homogenous and allowed to incubate for 3 minutes at room temperature. The tubes were then centrifuged at 12000 x g for 15 minutes at 4°C to achieve phase separation. Approximately 500 μl of the clear upper aqueous layer containing RNA was then transferred to a clean 1.5 mL tube and cold 70% ethanol was added in a 1:1 ratio. The samples were mixed gently by pipetting until the mixture was homogenous. The RNA was then transferred to PureLink RNA Mini Kit extraction columns (Thermo Fisher) for binding, washing, and elution. The rest of the procedure was performed according to the manufacturer’s instructions. Briefly, <700 μl of the sample was transferred to the PureLink columns and centrifuged at 12000 x g for 30 seconds at room temperature. This step was repeated until the entire sample was processed. The samples were then washed with Wash Buffer. An additional on-column DNase step was performed following the instructions for the PureLink DNase Set (Thermo Fisher, Cat. No. 12185010). DNase was added to the columns and allowed to incubate for 10 minutes. After DNase digestion, the samples were washed twice with Wash Buffer II and the RNA was then eluted in 50 μl of RNase-free water. The quality and concentration of the RNA was then assessed using the Agilent RNA 6000 Nano Kit (Agilent, Cat. No. 5067-1511) according to the manufacturer’s instructions.

#### cDNA synthesis

Total RNA was converted to cDNA using the SuperScript III First-Strand Synthesis System (Thermo Fisher, Cat. No. 18080051). For cDNA synthesis, a mixture of 1 μL of 50 ng/μl of random hexamers, 10 mM of dNTP mix, and approximately 1 μg of RNA was prepared before adding 10 μl of cDNA Synthesis Mix.

#### Primers

Two primers sets were used to test each stool sample for SARS-CoV-2 presence. The first primer set (probe-based) was taken directly from the CDC SARS-CoV-2 (2019-nCoV) RUO Primers and Probes (Integrated DNA Technologies, Cat. No. 10006713). The CDC 2019-nCoV positive plasmid control was also used (Integrated DNA Technologies, Cat. No. 10006625). Human RNaseP was used as a host control. The second primer set (dye-based) consisted of nsp14 primers targeting the SARS-CoV-2 exonuclease[45], bacterial 16S V4 primers for assessing the quality of RNA extracted from stool, and three human housekeeping gene controls targeting: hypoxanthine-guanine phosphoribosyltransferase (HPRT; low copy number gene), glyceraldehyde 3-phosphate dehydrogenase (GAPDH; mid copy number gene), and B-actin (high copy number gene). These various human housekeeping gene primers provided a comprehensive quantification of the mount of host DNA in feces, which was often low. Primer sequences are provided in Table S4.

#### qPCR

Each stool sample was tested for SARS-CoV-2 presence using both a probe-based and dye-based qPCR. The probe-based qPCR was performed using the Luna Universal Probe qPCR Master Mix (New England BioLabs, Cat. No. M3004L) and the CDC primers. Each reaction consisted of a total volume of 20 μl with 1.5 μl of 6.7 μM primer/probe mix, 4 μl of cDNA, 4.5 μl of water, and 10 μl of Luna Universal Probe qPCR Master Mix. The qPCR was carried out following manufacturer recommendations on a Viia7 Real-Time PCR system (Thermofisher). The dye-based qPCR was performed using the Luna Universal qPCR Master Mix (New England BioLabs, Cat. No. M3003L) and the remaining primers: nsp14, 16S, HPRT, β-actin, GAPDH. Each reaction consisted of a total volume of 20 μl with 1 μl of 10 μM of forward and reverse primers, 4 μl of cDNA, 5 μl of water, and 10 μl of Luna Universal qPCR Master Mix (NEB). The qPCR was carried out following manufacturer recommendation on a Viia7 Real-Time PCR system (Thermofisher).

### Measurement of fecal cytokines, calprotectin and total IgA

Fecal cytokines were assayed by sandwich ELISA (DuoSet® ELISA kits, R&D systems (IL-1β, IL-1ra, IL-6, IL-8, IL-10, IL-17 and TNFα or ThermoFisher Human ELISA kit (IL-23)) following the manufacturer’s instructions. For analysis of IL-23, fecal supernatants were diluted 1:2 in the kit-provided Sample Diluent A. For IL-1β and IL-1ra, fecal supernatants were diluted 1:5 in PBS with 1% BSA and for the remaining cytokines fecal supernatants were diluted 1:2 in PBS with 1% BSA. For measurement of fecal calprotectin, frozen heat-inactivated stool aliquots were further diluted in extraction buffer (BULHLMANN, EK-CAL2) and fecal Calprotectin levels were determined using the fCAL ELISA kit (BULHLMANN, EK-CAL2) according to manufacturer’s instructions. Total IgA was measured by ELISA using anti-IgA capture (1μg/mL; Southern Cat #2050-01) followed by anti-human IgA-HRP (1:8000, Southern Cat #2053-05) as previously described[46].

To comply with local biosafety protocols, it was necessary to treat samples to inactivate any potentially infectious virus present in the feces. We found that heating samples to 60°C for 60 minutes, a commonly used method for inactivation [32], led to degradation of recombinant cytokines spiked into stool samples. Some cytokines, including IL-1β, were particularly sensitive (Fig. S5). In contrast, 0.75% Triton X-100 included in the homogenization buffer (to achieve a final concentration of 0.5% when fecal samples were suspended 1:2 w:v), which is known to inactivate SARS-CoV-2 [44], and did not interfere with these assays. This pretreatment was used for all samples where proteins were measured (Fig. S5).

### ELISA for fecal SARS-CoV-2-specific IgA

The protocol was adapted from a published method [32, 37]. Polystyrene 96 half-area plates (Corning# 3690) were coated with 60μL of diluted SARS-CoV-2 RBD or S1 protein (2μg/mL in 1x PBS, produced in the laboratory of Dr. Florian Krammer) and incubated at 4°C overnight. Plates were washed with PBS plus 0.05% Tween20 (PBS-T) and blocked with PBS-T with 3% (w:v) milk powder (AmericanBio) for 2-3 hours. Fecal extract (pre-diluted 1:2 w:v, see above) or serum was mixed with equal volume PBS-T with 1% (w:v) milk powder and serially diluted two-fold for eight iterations. 30μL of the analyte dilution series was added to the plate for 2 hours. Plates were washed, and 60μL of anti-IgA-horseradish peroxidase (HRP) (Southern, 1:8000) or anti-mouse IgA (Millipore Sigma, 1:8000) was added. After 2 hours, ELISA plates were again washed and developed using TMB substrate (Biolegend). Plates were quenched with 2N sulfuric acid and read at 450nm. A titrated serum sample from a seropositive donor was included on every assay plate and used to normalize data between plates, correcting for minor batch variation.

### 16S and metagenomic microbiome sequencing

DNA extraction buffer (4.3 mM Tris, pH 8; 0.4 mM EDTA, 43 mM NaCl, 3% v/w SDS, 42% v/v phenol/chloroform/IAA and 21% v/v QIAquick PM buffer) containing 400 μl 0.1 mm Zirconia/Silica Beads was added to fecal samples and the samples were bead-beat on a BioSpec Mini-Beadbeater-96 for 2 minutes. After centrifugation (5 min, 4000 rpm), the ∼400 μl of aqueous layer was mixed with 650 μl QIAquick PM buffer DNA was purified using the QIAquick 96 PCR Purification Kit (Qiagen) according to manufactures protocol. DNA concentration was quantified using the Quant-iT dsDNA Assay Kit, broad range (Life Technologies) and normalized to 2 ng/μl on a Beckman handling robot. Amplicon preparation and sequencing was performed as previously described [47]. Briefly, bacterial 16S rDNA PCR including no template controls were setup in a separate PCR workstation using dual-indexed primers. PCR reactions contained 1 µM for each primer, 4 ng DNA, and Phusion Flash High-Fidelity PCR Master Mix (Thermo Fisher Scientific). Reactions were held at 98°C for 30 s, proceeding to 25 cycles at 98°C for 10 s, 45°C for 30 s, and 72°C for 30 s and a final extension of 2 min at 72°C. Amplicons were evaluated by gel electrophoresis. The sequencing library was prepared by combining equivolume amounts of each amplicon, size-selected and concentrated using AMPure XP beads (0.8X, Beckman). Library concentration was quantified by Qubit and qPCR, mixed with 15% PhiX, diluted to 4 pM and subjected to paired-end sequencing (Reagent Kit V2, 2×150bp) on an Illumina MiSeq sequencer. Resulting fastq files were analyzed using QIIME2 version 2019.10[48] and the DADA2[49] denoised-paired plugin with a truncation length of 150bp and 145bp for the forward and reverse read, respectively. Amplicon Sequence Variants (ASVs) were classified using the Scikit-Learn plugin [50] using the Naive Bayes classifiers trained on the silva-132-99-515-806-nb-classifier. Resulting ASV tables were filtered using a minimum depth of 5000, a minimal ASV frequency of 10 and minimal sample frequency of 2 and core metrics were calculated using the Qiime2 core metrics plugin.

Metagenomic sequencing was performed as previously described [51]. Briefly, DNA (extracted as for 16S rRNA amplicon sequencing) was sonicated and Illumina sequencing libraries generated using the NEBNext Ultra II DNA Library Prep Kit. Ligation products of 500– 600 base pairs were purified using SPRIselect beads (Beckman Coulter) and enrichment PCR performed. Samples were pooled in equal proportions and size-selected using AMPure XP beads (Beckman Coulter) before sequencing with an Illumina HiSeq (paired-end 150 bp). One sample yielding less than 100,000 reads was excluded from further analysis. Remaining samples were sequenced to an average depth of 1.52 × 10^6^ reads (SD 3.73 × 10^5^) per sample. Reads were trimmed with Trimmomatic [52] and taxonomic assignments were generated with MetaPhlAn2^[53]^. Data is deposited to NCBI under BioProject PRJNA660883.

## Viral Genomes

Complete viral genomes were generated as previously described[24] Briefly, cDNA was amplified using two sets of tiling PCR primers [54]. cDNA libraries were prepared with the Nextera ST DNA Sample preparation kit (Illumina) and sequenced on a MiSeq instrument (Illumina) on a 150 nt paired-ended configuration. Assembly was performed as described [24] using a custom pipeline (https://github.com/mjsull/COVID_pipe). To infer the divergence and phylogenetic relationships of the three genomes sequenced from stool samples relative to other global isolates, we used 403 viral genomes from nasopharyngeal samples from the Mount Sinai Hospital collected between February 29 and July 15, and a global background of 4,644 SARS-CoV-2 genomes available in GISAID EpiCov database [55]. Phylogenetic analysis was performed using Nextstrain build for SARS-CoV-2 genomes with default parameters[56]. The mutation profiles were analyzed with the tool NextClade v0.3.6 (https://clades.nextstrain.org/). Data is deposited to GISAID and Genbank.

## Data Availability

16S amplicon and metagenomic sequencing data has been deposited to NCBI under Bioproject PRJNA660883. Viral genomes have been deposited to GISAID and Genbank. De-identified raw source data used to generate all figures is provided in the supplemental information.

## Acknowledgments

We thank the study participants for their contributions to this study. We thank Joan Shang (Icahn School of Medicine at Mount Sinai) for assistance with assay optimization and India Brough, Ciara Latore, Emily Thornton, Fiona Powrie and Paul Bowness (University of Oxford) for helpful discussions about measuring cytokines in fecal samples. We thank the authors from originating and submitting laboratories of sequences from GISAID EpiCov (www.gisaid.org) that were used as background for the phylogenetic analysis. Funding: This work was supported by grants from the NIH (NIGMS GM108505, NIDDK DK112978, and NIDDK DK124133 to J.J.F and NIDDK DK123749 to S.M. and J.J.F.). G.J.B. is supported by a Research Fellowship Award from the Crohn’s and Colitis Foundation of America. M.P.S. is supported by NIH T32 5T32AI007605. H.v.B. and A.S.G.-R were supported in part by the Centers of Excellence for Influenza Research and Surveillance contract HHSN272201400008C and A.S.G.-R. was further supported in part by a Robin Chemers Neustein Postdoctoral Fellowship Award. Work in the Krammer laboratory was partially supported by the NIAID Centers of Excellence for Influenza Research and Surveillance (CEIRS) contract HHSN272201400008C, Collaborative Influenza Vaccine Innovation Centers (CIVIC) contract 75N93019C00051, and the generous support of the JPB foundation, the Open Philanthropy Project (#2020-215611) and other philanthropic donations. Computational resources used in this research were further supported by the Office of Research Infrastructure of the National Institutes of Health (NIH) under awards S10OD018522 and S10OD026880.

## Author contributions

G.J.B.; study concept and design; acquisition of data; analysis and interpretation of data; statistical analysis; drafting of the manuscript, A.C-L.; study concept and design; acquisition of data; analysis and interpretation of data; statistical analysis; drafting of the manuscript, F.C.; study concept and design; acquisition of data; analysis and interpretation of data; statistical analysis; drafting of the manuscript, A.E.L.; study concept and design; acquisition of data; analysis and interpretation of data; statistical analysis; drafting of the manuscript, M.P.S.; study concept and design; acquisition of data; analysis and interpretation of data; statistical analysis; drafting of the manuscript, T.P.; acquisition of data, J.E.; acquisition of data, I.M.; acquisition of data, A.S.G-R.; acquisition of data; analysis and interpretation of data;, S.S.; acquisition of data, M.T.; acquisition of data, L.T.G.; acquisition of data, R.E.D; acquisition of data, D.J.; acquisition of data, A.v.d.G.; acquisition of data, Z.K.; acquisition of data, G.M-D.; acquisition of data, F.A.; technical, or material support;, D.A.H.; technical, or material support;, B.R.T.; technical, or material support, M.C.D.; technical, or material support;, M.M.; technical, or material support, H.v.B.; study supervision; technical, or material support;, F.K. study supervision; technical, or material support;, G.B.; acquisition of data; analysis and interpretation of data; statistical analysis; study concept and design, S.M.; study supervision; study concept and design; analysis and interpretation of data;, J.J.F.; study supervision; study concept and design; analysis and interpretation of data.

## Declaration of interests

M.C.D is a consultant for Abbvie, Arena, BMS, Boehringer Ingelheim, Genentech, Janssen, Pfizer, Prometheus Biosciences, Takeda, Target RWE and UCB, has received research support from Janssen, Pfizer, Prometheus Biosciences and Abbvie and is co-founder of Cornerstones Health, Mitest Health, Trellus Health, M.M. is a consultant for Takeda, Genentech, Regeneron, Compugen, Myeloid Therapeutics. F.K. declares that The Icahn School of Medicine at Mount Sinai has filed patent applications regarding SARS-CoV-2 serology assays and has licensed reagents to several commercial entities. J.J.F. serves on the scientific advisor board of Vedanta Biosciences. The remaining authors declare no competing of interest.

## Data availability

Microbiome sequencing data is deposited to NCBI under BioProject PRJNA660883. Viral genomes are deposited in GISAID and accession numbers are provided in Data File S2. All the deidentified clinical and assay data used to generate the figures in this manuscript are provided in Date files S1-3.

## Notes

### Author Declarations

The study was reviewed and approved by the institutional review board at Icahn School of Medicine at Mount Sinai (Protocol #: HS# 16-00512/ GCO# 16-0583 and HS# 11-01669).

### Summary of Updates

We have updated the manuscript to include analysis of SARS-CoV-2 genomes isolated from fecal samples.

