## Supplemental Figures for "Limited intestinal inflammation despite diarrhea, fecal viral RNA and SARS-CoV-2-specific IgA in patients with acute COVID-19"

**Supplementary Materials:**

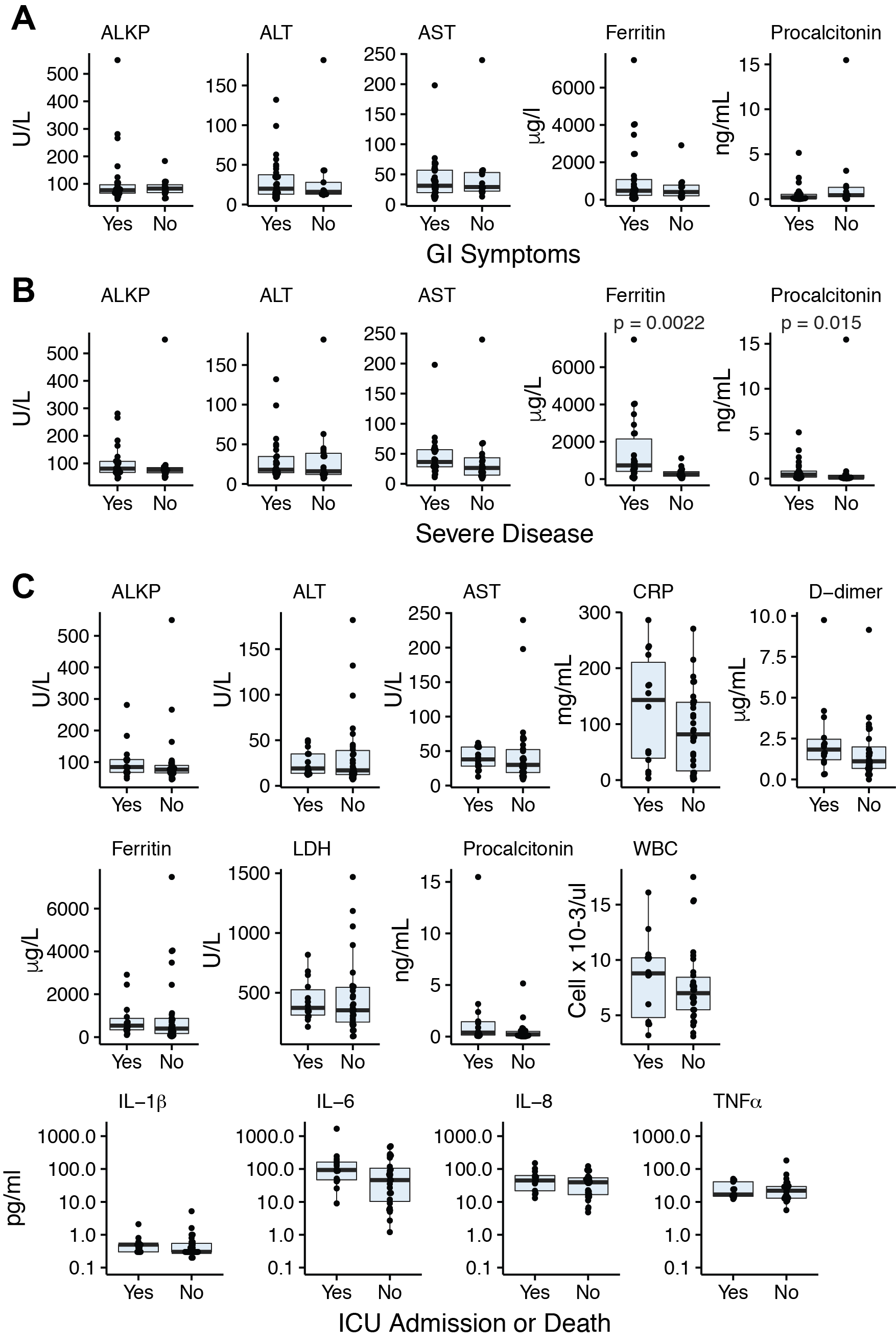

**Fig. S1. Gastrointestinal symptoms and serologic parameters in hospitalized COVID-19 patients**. **(A)** Liver enzymes and laboratory values in hospitalized COVID-19 patients with and without GI symptoms. **(B)** Liver enzymes and laboratory values in hospitalized COVID-19 patients with severe disease or non-severe disease. Each point represents an individual value for a patient, the box plot shows the median and the interquartile range and the p-values are calculated using the Mann-Whitney test with significance defined as p<0.05.

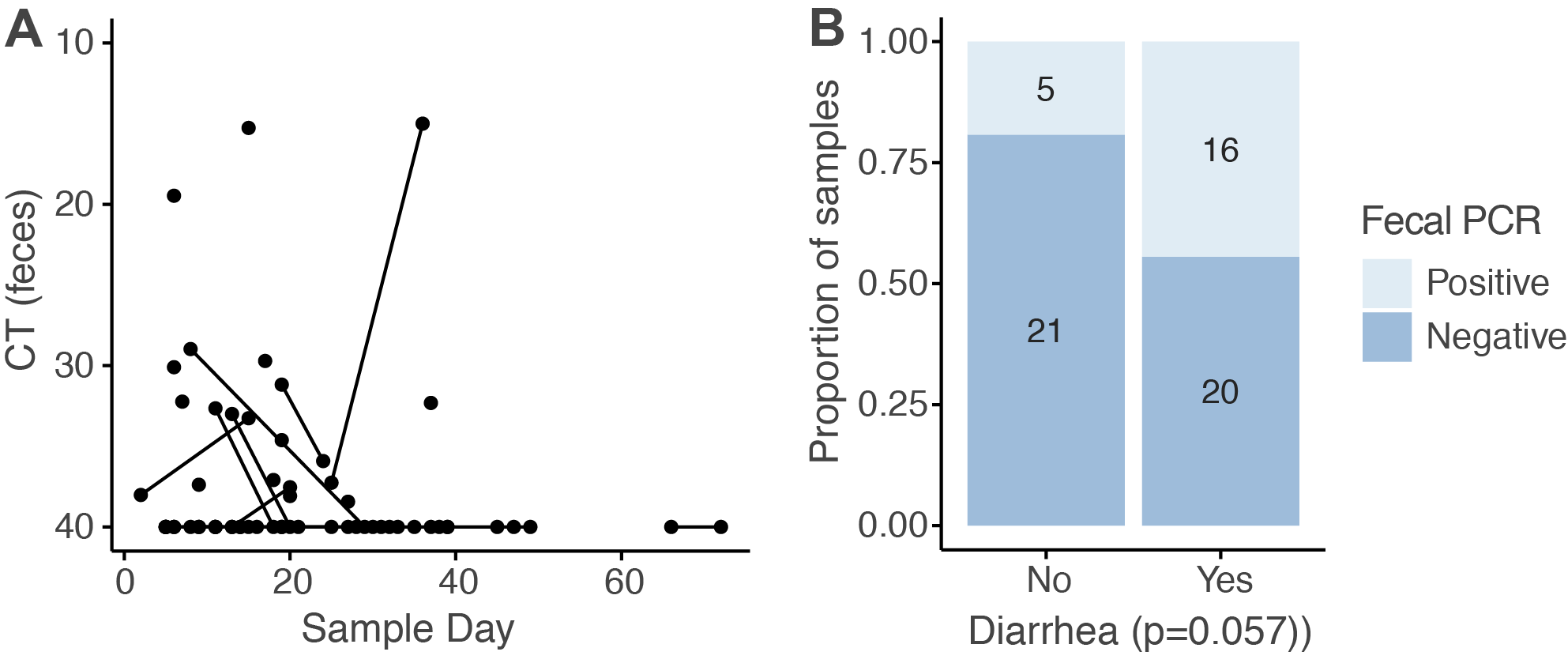

**Figure S2.** **Detection of SARS-CoV-2 virus genome in stool. (A)**. Each point shows the median SARS-CoV-2 Ct from one sample, plotted against the sampling day relative to the onset of symptoms. Lines connect samples from the same donor. (**B**) The proportion of positive and negative fecal SARS-CoV-2 PCR results from donors with and without diarrhea. p value - Fisher's exact test.

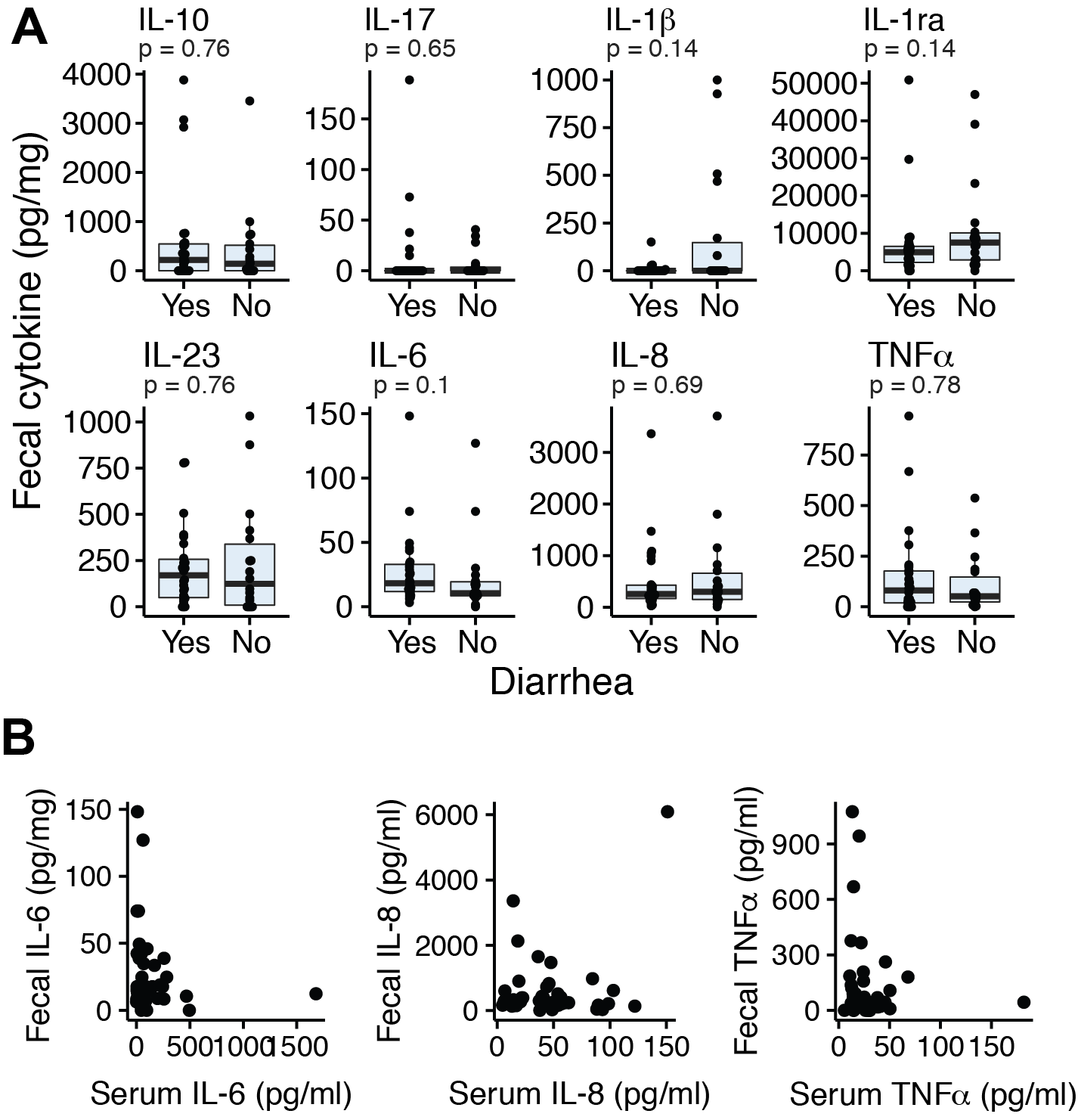

**Fig. S3. Fecal cytokine levels in COVID-19 patients.** (**A**) Concentrations of the indicated cytokine in fecal samples from COVID-19 patients with and without diarrhea. P values - Mann-Whitney. **(B)** Relationship between concentrations of the indicated cytokine in feces and serum.

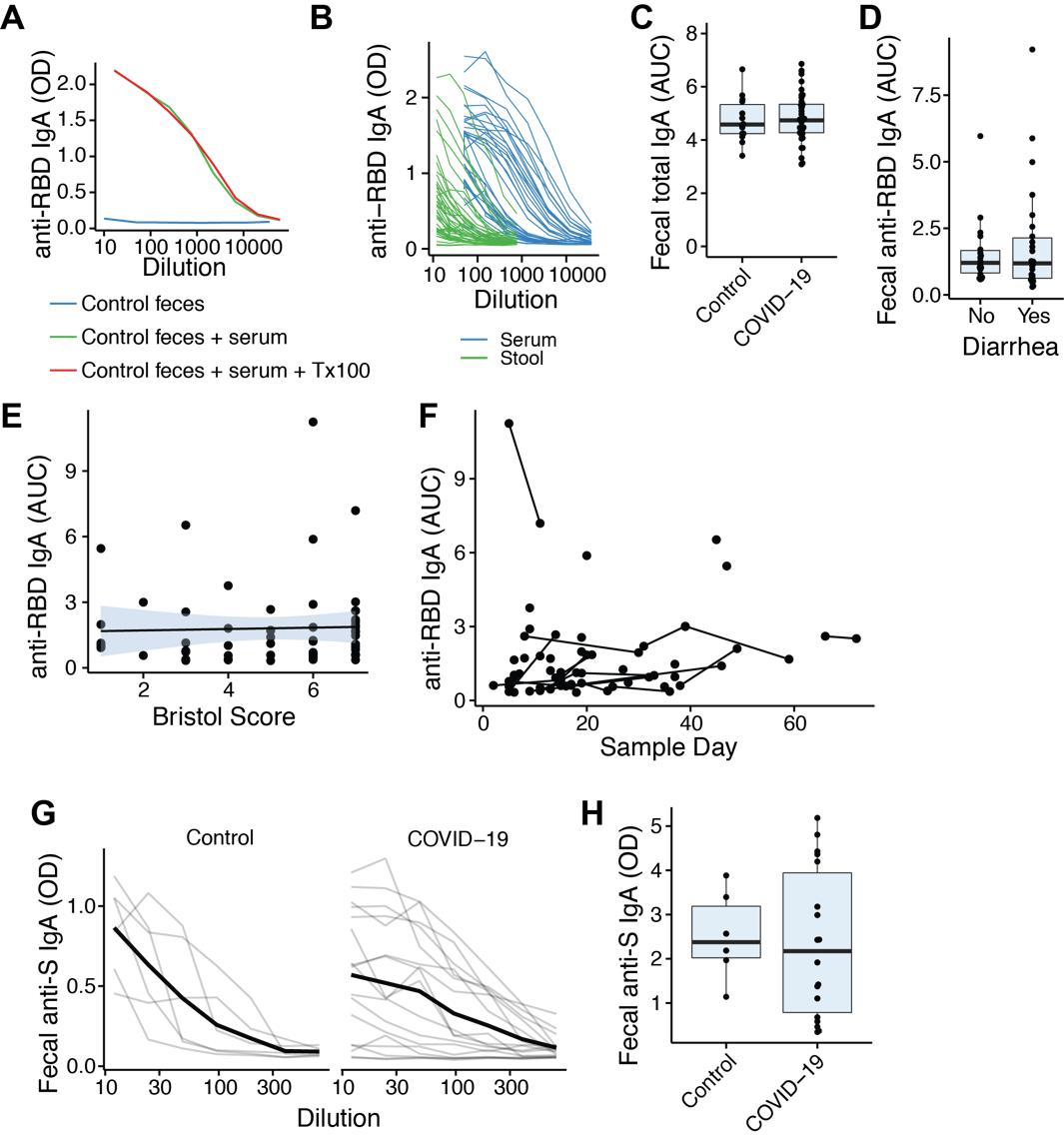

**Figure. S4.** **SARS-CoV-2-specific IgA in stool of COVID-19 patients. (A**) Triton X-100 does not reduce detection of anti-RBD IgA in a serum sample from a seropositive donor serum spiked into a healthy donor stool sample. **(B)** Relative titers of serum and fecal anti-RBD IgA in samples from COVID-19 patients. **(C)** Relative titers of total IgA in feces of control donors and COVID-19 patients. (**D** and **E)** Anti-SARS-CoV-2 RBD specific IgA titers are not different in donors with diarrhea. (D – averaged by donor, E - by sample). **(F)** Anti-SARS-CoV-2 RBD IgA in feces over time relative to reported symptom onset. Each point shows data from one sample and lines connect samples from the same donor. (**G**) Anti-SARS-CoV-2 Spike protein IgA in control donors and COVID-19 patients. (**H**) Relative titers of anti-SARS-CoV-2 Spike protein IgA in control donors and COVID-19 patients.

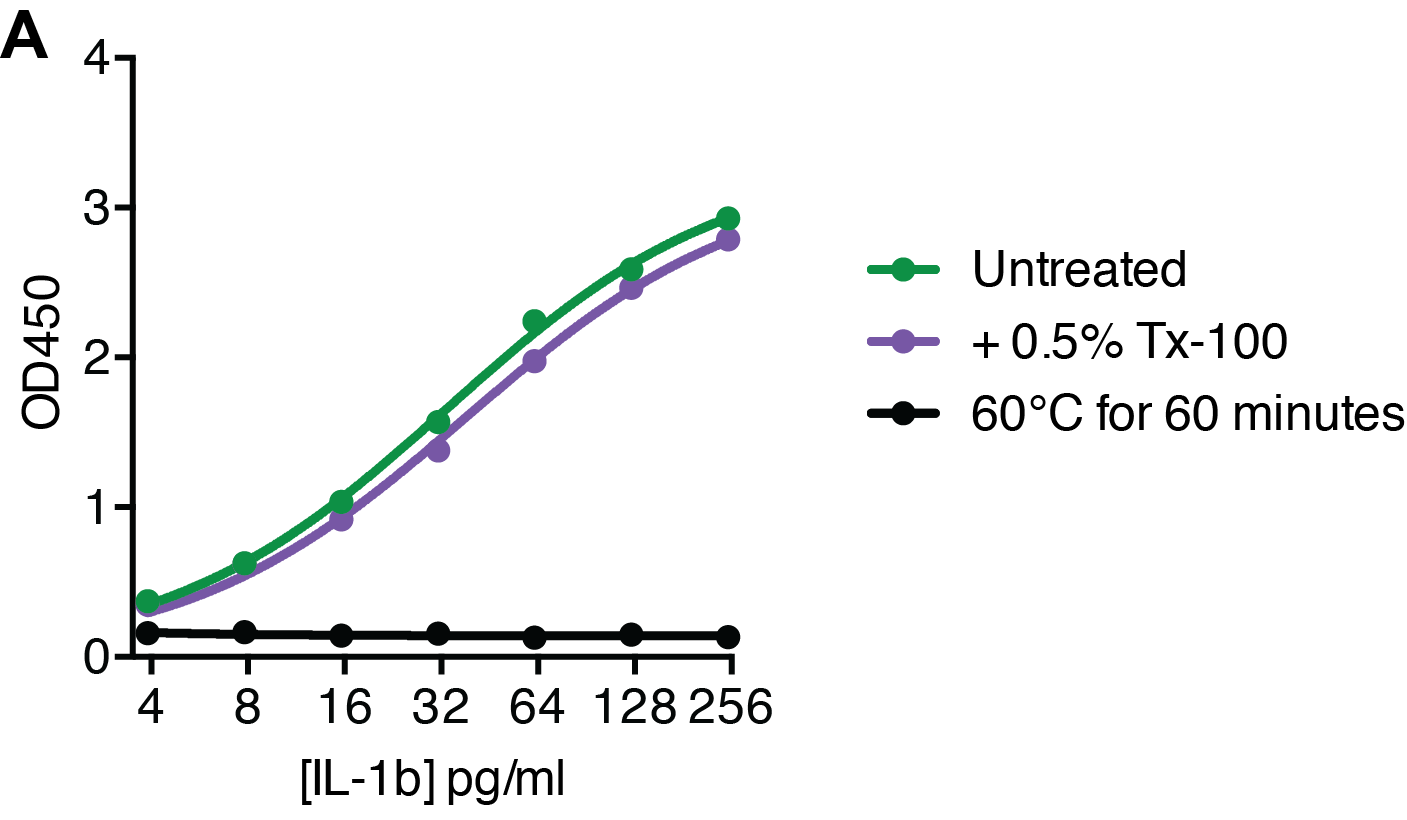

**Figure S5. Virus inactivation method compatible with cytokine ELISAs. (A)** Recombinant human IL-1β was spiked into a stool homogenate from a healthy donor and either heated at 60°C for 60 minutes or treated with 0.5% Triton X-10 (Tx-100) for 60 minutes at 4C before ELISA analysis.

| **Table S1: Sample collection and description**  *The number of each sample type and the day of collection (median +/- the range), relative to the onset of COVID-19 symptoms.* | | | | |
| --- | --- | --- | --- | --- |
|  | 1st stool sample | 2nd stool sample | | Serum |
| Number of samples | 44 | 18 | | 32 |
| Median time in days (range) from symptom onset to sample collection | 16 (2-66) | 24.5 (11-72) | | 16 (3-46) |

| **Table S2: COVID-19 treatments in patients with and without GI symptoms**  *The absolute number and percentage of the cohort (in parentheses) who received the indicated therapeutic. Statistical comparisons are by Fisher’s exact test.* | | | | | | | |
| --- | --- | --- | --- | --- | --- | --- | --- |
|  | Total  (n=44) | | GI symptoms (n=31) | | No GI symptoms (n=13) | | p-value |
| Antibiotics | 28 (63.6) | | 17 (54.8) | | 11 (84.6) | | 0.09 |
| Azithromycin | 17 (38.6) | | 8 (25.8) | | 9 (69.2) | | **0.02** |
| Vancomycin | 13 (29.5) | | 7 (22.6) | | 6 (46.2) | | 0.16 |
| Ceftriaxone (CTX) | 17 (38.6) | | 9 (29.0) | | 8 (61.5) | | 0.09 |
| Vancomycin + CTX | 8 (18.2) | | 4 (12.9) | | 4 (30.8) | | 0.21 |
| Cefepime | 12 (27.3) | | 8 (25.8) | | 4 (30.8) | | 0.73 |
| Other antibiotics | 10 (22.7) | | 5 (16.1) | | 5 (38.5) | | 0.13 |
| Plasma | 20 (45.5) | | 13 (41.9) | | 7 (53.8) | | 0.52 |
| Plasma before  sample collection | 19 (43.2) | | 13 (41.9) | | 6 (46.2) | | >0.99 |
| Hydroxychloroquine | 19 (43.2) | | 12 (38.7) | | 7 (53.8) | | 0.51 |
| Remdesivir | 13 (29.5) | | 8 (25.8) | | 5 (38.5) | | 0.48 |
| Steroids | 18 (40.9) | | 13 (41.9) | | 5 (38.5) | | >0.99 |
| Therapeutic anticoagulation | 31 (70.5) | | 22 (71.0) | | 9 (69.2) | | >0.99 |

**Table S3: Disease severity associated with composite outcome of ICU admission or death**

*The absolute number and percentage of the cohort (in parentheses) with each indicated COIVD-19 severity. Statistical comparisons are by Fisher’s exact test.*

| **Severity on admission** | ICU admission or death (n=14) | Neither  (n=30) | p-value |
| --- | --- | --- | --- |
| Mild or moderate | 6 (42.9) | 22 (73.3) | 0.09 |
| Severe | 8 (57.1) | 8 (26.7) |  |
| **Peak severity** |  |  |  |
| Mild or moderate | 2 (14.3) | 16 (53.3) | **0.02** |
| Severe | 12 (85.7) | 14 (46.7) |  |

| **Table S4. PCR primer sequences** | | |
| --- | --- | --- |
| **Primer** | **Sequence (5’-3’)** | **Reference** |
| 2019-nCoV_N1 Forward | GACCCCAAAATCAGCGAAAT | [57] |
| 2019-nCoV_N1 Reverse | TCTGGTTACTGCCAGTTGAATCTG | [57] |
| 2019-nCoV_N1 Probe | ACCCCGCATTACGTTTGGTGGACC | [57] |
| 2019-nCoV_N2 Forward | TTACAAACATTGGCCGCAAA | [57] |
| 2019-nCoV_N2 Reverse | GCGCGACATTCCGAAGAA | [57] |
| 2019-nCoV_N2 Probe | ACAATTTGCCCCCAGCGCTTCAG | [57] |
| RNase P Forward | AGATTTGGACCTGCGAGCG | [57] |
| RNase P Reverse | GAGCGGCTGTCTCCACAAGT | [57] |
| RNase P Probe | TTCTGACCTGAAGGCTCTGCGCG | [57] |
| Nsp14 Forward | TGGGGYTTTACRGGTAACCT | [45] |
| Nsp14 Reverse | AACRCGCTTAACAAAGCACTC | [45] |
| Hypoxanthine phosphoribosyl transferase (HPRT) Forward | CAACAGGCTTTTCCTGGTT | [58] |
| Hypoxanthine phosphoribosyl transferase (HPRT) Reverse | GGCTACTCTGCCCATGAAGA | [58] |
| ß-actin Forward | CCCAGCACAATGAAGATCAA | [59] |
| ß-actin Reverse | ACATCTGCTGGAAGGTGGAC | [59] |
| Glyceraldehyde 3-phosphate dehydrogenase (GAPDH) Forward | GTCGTGGAGTCTACTGGTGTCTTC | [60] |
| Glyceraldehyde 3-phosphate dehydrogenase (GAPDH) Reverse | GTCATATTTCTCGTGGTTCACACC | [60] |
| 16S 8F (qPCR control) | AGAGTTTGATCCTGGCTCAG | [61] |
| 16S 1391R (qPCR control) | GACGGGCGGTGWGTRCA | [61] |

**Supplemental Text:**

Below is the clinical questionnaire administered to all recruited COVID-19 patients.

**Clinical Questionnaire:**

**Simple questionnaire:**

1. Diarrhea (Y/N)?
2. Nausea (Y/N)?
3. Vomiting (Y/N)?

**Detailed questionnaire:**

1. Are you having diarrhea? (Y/N)
2. When did your diarrhea start? (date)
3. When did any of your other symptoms start including fever or other symptoms (cough, shortness of breath, sore throat? (date)
4. Did your diarrhea end or is it ongoing?
5. How many stools are you having per day?
6. Is there any bood in your stool? (Y/N)
7. Are stools large volume and mostly water? (Y/N)
8. Are you waking up in the middle of the night to have bowel movement? (Y/N)
9. Do you have abdominal pain? (Y/N)

**Other GI symptoms / history question:**

1. Is your appetite decreased? (Y/N)
2. Are you having nausea? (Y/N)
3. Are you having any episodes of vomiting? (Y/N)
